# Early Cardiac Rehabilitation After Acute Heart Failure Hospitalization and Its Long-Term Benefits in the Presence of Cardiac Fibrosis: A Prospective Cohort Study

**DOI:** 10.1101/2025.04.28.25326610

**Authors:** Shyh-Ming Chen, Lin-Yi Wang, Hao-Yi Hsiao, Chin-Ling Wei, You-Cheng Zheng, Po-Jui Wu, Chien-Jen Chen, Chi-Ling Hang, Steve Leu, Yung-Lung Chen

## Abstract

**Aim:** To evaluate whether initiation of cardiac rehabilitation (CR) within 6 weeks improves long-term outcomes of patients with acute heart failure (HF).

**Methods:** Patients with acute HF who participated in an HF disease management program from January 2019 to July 2022 were prospectively enrolled. Eligible patients were divided into two groups: early CR (and continued home-based CR during the follow-up period) and non-CR. The primary outcome was all-cause mortality. Secondary outcomes were rehospitalisation for recurrent HF and changes in 12-item Kansas City Cardiomyopathy Questionnaire scores from baseline to 6 months and 1 year. A post-hoc analysis stratified by lysyl oxidase-like 2 (LOXL2) levels assessed CR benefits for patients with cardiac fibrosis.

**Results:** Of 162 patients, 34 received early CR. The non-CR group was older (median age: 58.5 vs. 53.0 years, p=0.022) and had higher N-terminal pro-B-type natriuretic peptide levels (4552.5 vs. 1275.0 pg/mL, p=0.002). Propensity score matching yielded 33 patients per group. Over 2.85 years, the early CR group had lower all-cause mortality (0 vs. 87.16 events per 1000 patient-years, rate difference: −0.087 [95% confidence interval {CI}: −0.143 to −0.031], p=0.002). Patients with LOXL2 >200 pg/mL benefited the most (0 vs. 172.3 events per 1000 patient-years, rate difference: −0.172 [95% CI: −0.299 to −0.046], p=0.008).

**Conclusion:** Early post-discharge exercise-based CR was associated with reduced all-cause mortality in patients with acute HF. Patients with more severe cardiac fibrosis, indicated by higher LOXL2 levels, derived greater benefits from the CR program. Large-scale trials are needed to validate these findings.

**Trial registration:** The study protocol was registered at ClinicalTrials.gov (identifier: NCT03782337).

**Lay summary:**

- Early exercise after discharge for patients with acute heart failure is feasible, and early cardiac rehabilitation with nearly 3 years of follow-up is associated with a reduction in all-cause mortality without significant risks.
- Furthermore, for patients with severe cardiac fibrosis, cardiac rehabilitation can result in an even greater reduction in all-cause mortality.

Based on our findings, cardiac rehabilitation and exercise recommendations should be initiated early after discharge in patients with acute heart failure, particularly those with cardiac fibrosis.

## Introduction

Owing to increased life expectancy and advancements in the treatment of myocardial infarction, heart failure (HF) has become a growing problem globally. A study using the Taiwan National Health Insurance Research Database indicated an increase in the prevalence of HF from 0.63% in 2001 to 1.40% in 2016, representing a 2.22-fold increase over 16 years^1^. Despite medical advancements, the clinical prognosis of HF has not improved proportionately, with the estimated incident mortality rates of newly diagnosed HF being 38.5% at 2 years and 75.5% at 10 years^1^. Thus, reducing mortality in acute HF and preventing rehospitalisation for HF are critical challenges in modern medicine.

Cardiac rehabilitation (CR) is widely recognised as an essential tool for the secondary prevention of HF. CR programs usually include organised exercise training, which has been consistently proven to improve exercise capacity in individuals with HF. Improved exercise tolerance is linked to better overall cardiovascular function, reduced symptoms, and enhanced quality of life^2^. The Heart Failure: A Controlled Trial Investigating Outcomes of Exercise Training (HF-ACTION) showed that aerobic exercise training improved the quality of life in patients with HF with reduced ejection fraction (HFrEF) and that after prespecified adjustment, exercise training appeared to be associated with a reduction in cardiovascular mortality or hospitalisation for HF^3^. The HF-ACTION provided evidence supporting CR for stable chronic HF in patients with HFrEF categorised as New York Heart Association (NYHA) classes II–IV despite being on optimal therapy for ≥6 weeks. A retrospective study also suggested that early CR (at <6 weeks after discharge) might have beneficial effects on patients with acute HF who had just been discharged^4^. The Rehabilitation Therapy in Older Acute Heart Failure Patients (REHAB-HF) trial is a multicentre, single-blind randomised controlled trial (RCT) that assessed the effect of exercise training in old patients hospitalised for acute HF. In the REHAB-HF trial, the intervention was initiated during or early after hospitalisation and continued for phase II CR after discharge. The primary endpoint was the Short Physical Performance Battery (SPPB) scores at 3 months. The results indicated least-squares mean (±standard error) SPPB scores of 8.3±0.2 in the intervention group and 6.9±0.2 in the control group at 3 months (mean between-group difference: 1.5, 95% confidence interval [CI]: 0.9–2.0, *p*<0.001)^5^. Current evidence remains insufficient to definitively assess the impact of early rehabilitation on clinical hard endpoints in patients with acute HF, and the long-term effects of early rehabilitation in patients with acute HF have not been fully investigated.

The presence and extent of cardiac fibrosis substantially affect the prognosis of HF, as it can lead to contractile dysfunction and arrhythmic events^6^. Cardiac fibrosis is considered a key therapeutic target in HF management^7^. Nevertheless, existing therapies aimed at reversing cardiac fibrosis generally focus on nonspecific aspects of the fibrotic process, and the resilience of fibrosis to treatment poses a challenge in controlling fibrotic remodelling^8^. The potential benefits of early exercise training on cardiac fibrosis have been rarely studied in human participants and therefore warrant further investigation.

The current prospective cohort-controlled study aimed to evaluate whether early initiation of CR within 6 weeks could improve the long-term outcomes in patients with acute HF. We hypothesised that early multidisciplinary CR may improve the long-term survival rates in patients with acute HF compared with those in patients not undergoing CR and that patients with cardiac fibrosis may benefit from CR.

## Methods

### Study design and participants

This study enrolled patients who were hospitalised for acute HF at a referral medical centre. All patients were classified to have NYHA classes II–IV and had B-type natriuretic peptide (BNP) levels ≥100 pg/mL or N-terminal pro-BNP (NT-proBNP) levels ≥400 pg/mL (≥900 pg/mL in cases of concomitant atrial fibrillation). The inclusion criteria were as follows: patients (i) who were ≥20 years of age, (ii) experienced acute HF from January 2019 to July 2022, (iii) had a left ventricular ejection fraction (LVEF) of 40% or less, (iv) completed an HF Disease Management Program (HFDMP), and (v) were discharged alive. The exclusion criteria were as follows: an estimated survival time of <6 months, long-term bedridden status for >3 months, inability to tolerate an exercise test because of musculoskeletal disorders, inability to cooperate with functional assessments, ventilator dependence, terminal HF, refusal to participate by the family, and presence of severe primary valvular heart disease.

Eligible patients were divided into two groups according to their participation in outpatient exercise-based phase II CR and continuation of a home-based exercise program. An HF specialist nurse contacted the patients via phone within 1 week after discharge and then every 6 months until 31 July 2023. Patients were referred to outpatient rehabilitation clinics within 1 week after discharge. For patients who were willing to participate, exercise-based CR was scheduled within 6 weeks. The HF specialist nurse recorded each CR session attended. The home-based exercise program was designed as a continuation of the principles taught during phase II CR and included a mixture of aerobic, strength, and flexibility exercises tailored to each patient’s health status and abilities. An exercise physical therapist provided written and visual materials to guide patients’ routines at home. The HF specialist nurse monitored the effectiveness of both centre-based and home-based CR programs to ensure optimal outcomes.

This study was conducted in accordance with the principles embodied in the Declaration of Helsinki and was approved by the Institutional Review Board (IRB) of Chang Gung Medical Foundation (IRB approval number: 201801077B0C506). Written informed consent was obtained from all patients prior to enrolment. The study protocol was registered at ClinicalTrials.gov (identifier: NCT03782337).

### Exercise-based CR

Prior to discharge, all patients received the HFDMP, which included psychological assessment; education by an HF specialist nurse; and consultations with a dietitian, physiatrist, and psychologist^9,10^. Patients were encouraged to begin phase II CR within 6 weeks after discharge. Individualised continuous moderate-intensity aerobic exercise training was prescribed on the basis of cardiopulmonary exercise test results^11^. The training intensity was set within 10 beats of the anaerobic threshold or at 40%–60% of peak VO_2_, with adjustments being made every 2 weeks as tolerated (Borg scale of 12–14). Patients who completed at least one exercise session of phase II CR and continued the home-based exercise program were considered to have undergone CR.

### Blood samples and lysyl oxidase-like 2 (LOXL2) measurements

Fasting blood samples (≤10 mL) were obtained from patients after overnight fasting. The samples were centrifuged and stored at −70°C until the analysis of serum LOXL2 levels. Serum LOXL2 concentrations were measured using ELISA kits (R&D Systems, Abingdon, UK) according to the manufacturer’s protocol. Absorbance was measured at 450 nm using a microplate reader, and concentrations were quantified using corresponding standard curves.

### Outcomes

The primary outcome was all-cause mortality during the follow-up period. The secondary outcomes were recurrent rehospitalisation for HF and changes in the 12-item Kansas City Cardiomyopathy Questionnaire (KCCQ-12) scores from baseline to 6 months and 1 year.

### Statistical analysis

Sample size was calculated based on our previous study^4^. A minimum of 126 participants evenly distributed between the control and intervention groups were required for this study. To achieve 80.3% power with a significance level of 0.050, a two-sided log-rank test was performed, which detected a hazard ratio of 0.2319, assuming 3-year survival rates of 74.5% in the control group and 93.4% in the intervention group. This study was conducted over four time periods, with the participants being enrolled during the first three periods.

The normality of variable distributions was assessed using the Kolmogorov– Smirnov test. Patient demographic data were compared using Pearson’s chi-square test or Fisher’s exact test for categorical variables and the Mann–Whitney *U* test or Wilcoxon *W* test for continuous variables, with the results presented as medians and interquartile ranges (IQRs). A propensity score was calculated by fitting a logistic regression model after adjustment for factors such as age, sex, ischaemic cardiomyopathy, diabetes mellitus, hypertension, hyperlipidaemia, prior myocardial infarction, atrial fibrillation, body mass index, use of more than three types of guideline-directed medical therapy, estimated glomerular filtration rate, and LOXL2 levels.

A 1:1 nearest-neighbour matching without replacement was performed between the early CR and non-CR groups. Covariate balance before and after matching was assessed by comparing the standardised mean difference (SMD), with SMD <0.1 denoting a negligible imbalance between the two groups. Clinical outcomes, including all-cause mortality, rehospitalisation for HF, and improvements in KCCQ-12 scores at 6 and 12 months after propensity score matching (PSM), were compared between patients with and without CR using Pearson’s chi-square or Fisher’s exact test and the Mann–Whitney *U* test.

For the analysis of associations between CR and outcomes, incidence rates per 1000 patient-years and incidence rate differences were calculated for all-cause mortality and rehospitalisation for HF after PSM. Actuarial survival rates were estimated using the Kaplan–Meier method, and between-group differences were statistically examined using the log-rank test. A post-hoc analysis was conducted to explore whether CR improved the prognosis related to cardiac fibrosis. Patients were categorised into three groups according to LOXL2 levels: 0–100 pg/mL, >100 to ≤200 pg/mL, and >200 pg/mL. Additionally, incidence rates per 1000 patient-years and incidence rate differences for all-cause mortality were calculated to analyse the association between CR and outcomes, stratified by LOXL2 levels at admission. All statistical analyses were performed using SPSS software version 25.0 (IBM Corp., Armonk, NY, USA), with statistical significance set at *p*<0.05.

## Results

### Patient characteristics

A total of 480 patients with acute HF participated in the HFDMP from January 2019 to July 2022. Among them, 162 patients met both the inclusion and exclusion criteria and agreed to participate in this study. Overall, the early CR group comprised 34 patients who participated in the phase II exercise training program and continued to undergo home-based CR. In the CR group, 34 patients participated in centre-based CR, with a median of 7.0 rehabilitation exercise sessions (IQR: 1.00–27.25). PSM resulted in 33 patients per group (*Figure 1*).

**Figure 1.**
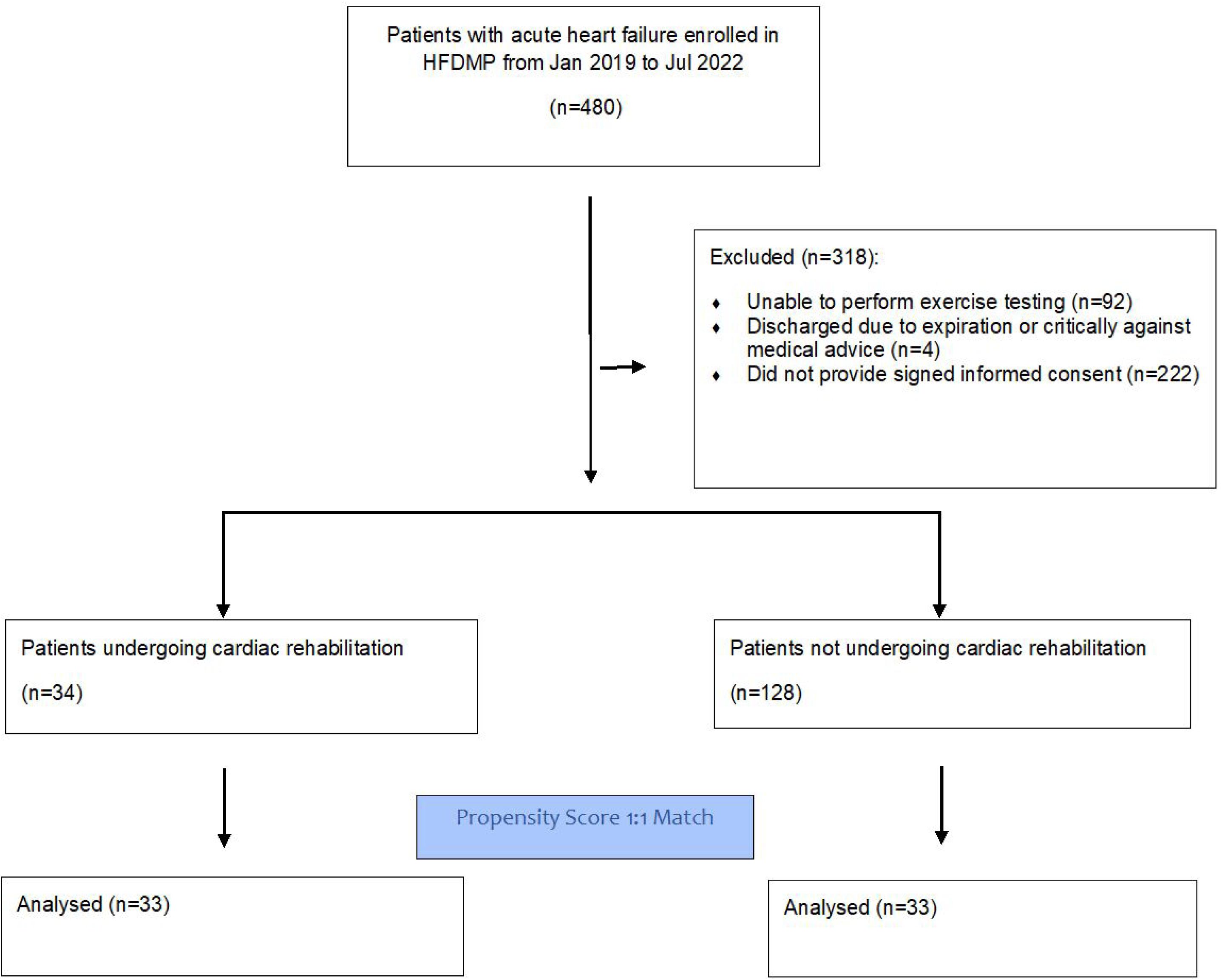
Flowchart of the study, HFDMP, Heart Failure Disease Management Program

Before PSM, patients in the non-CR group were older than those in the early CR group (median age: 58.5 vs. 53.0 years, *p*=0.022) and exhibited higher NT-proBNP levels (median: 4552.5 vs. 1275.0 pg/mL, *p*=0.002) (*Table 1*). After PSM, the variables were well-balanced between the two groups (*Table 2*).

**Table 1.**
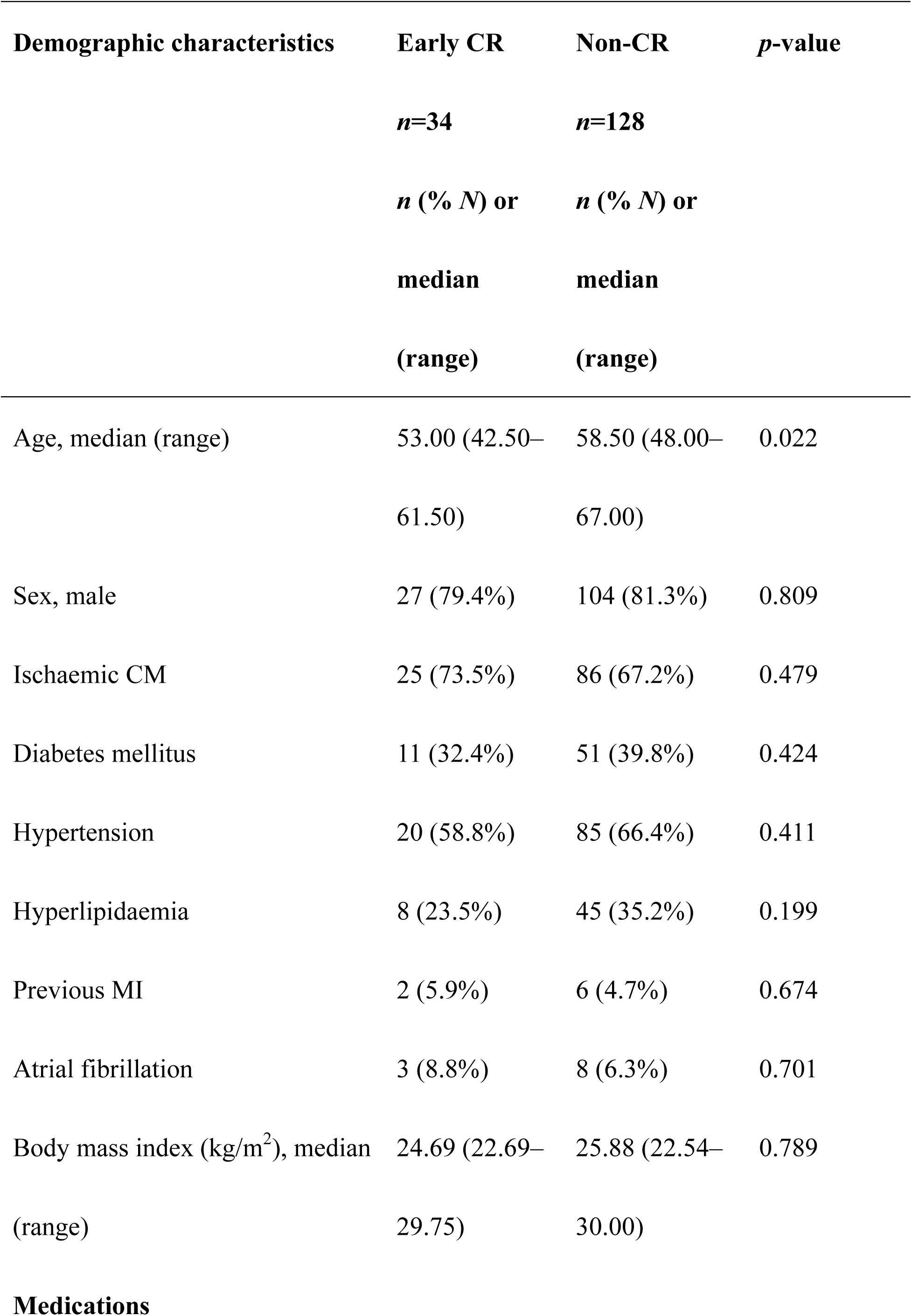
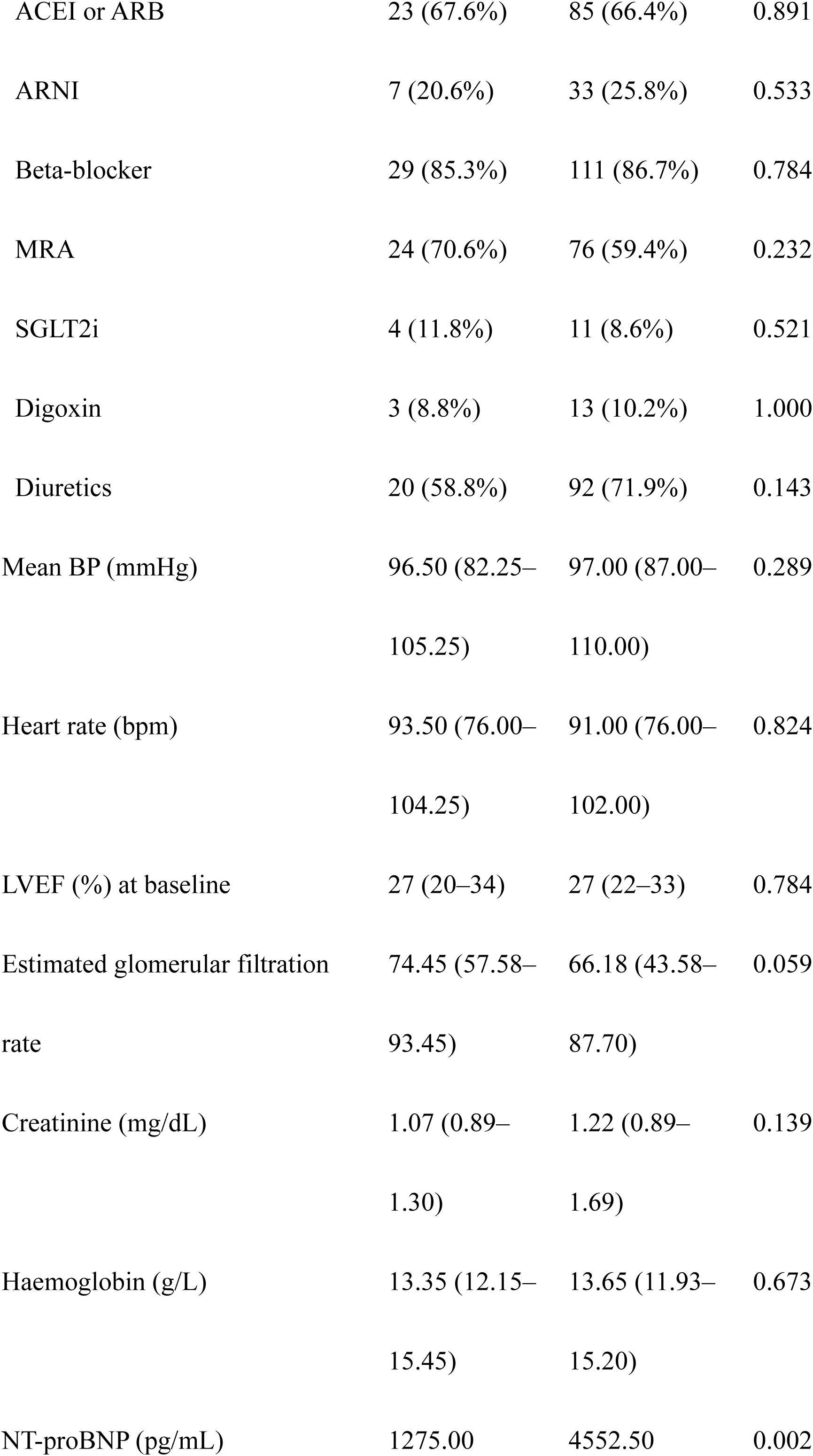
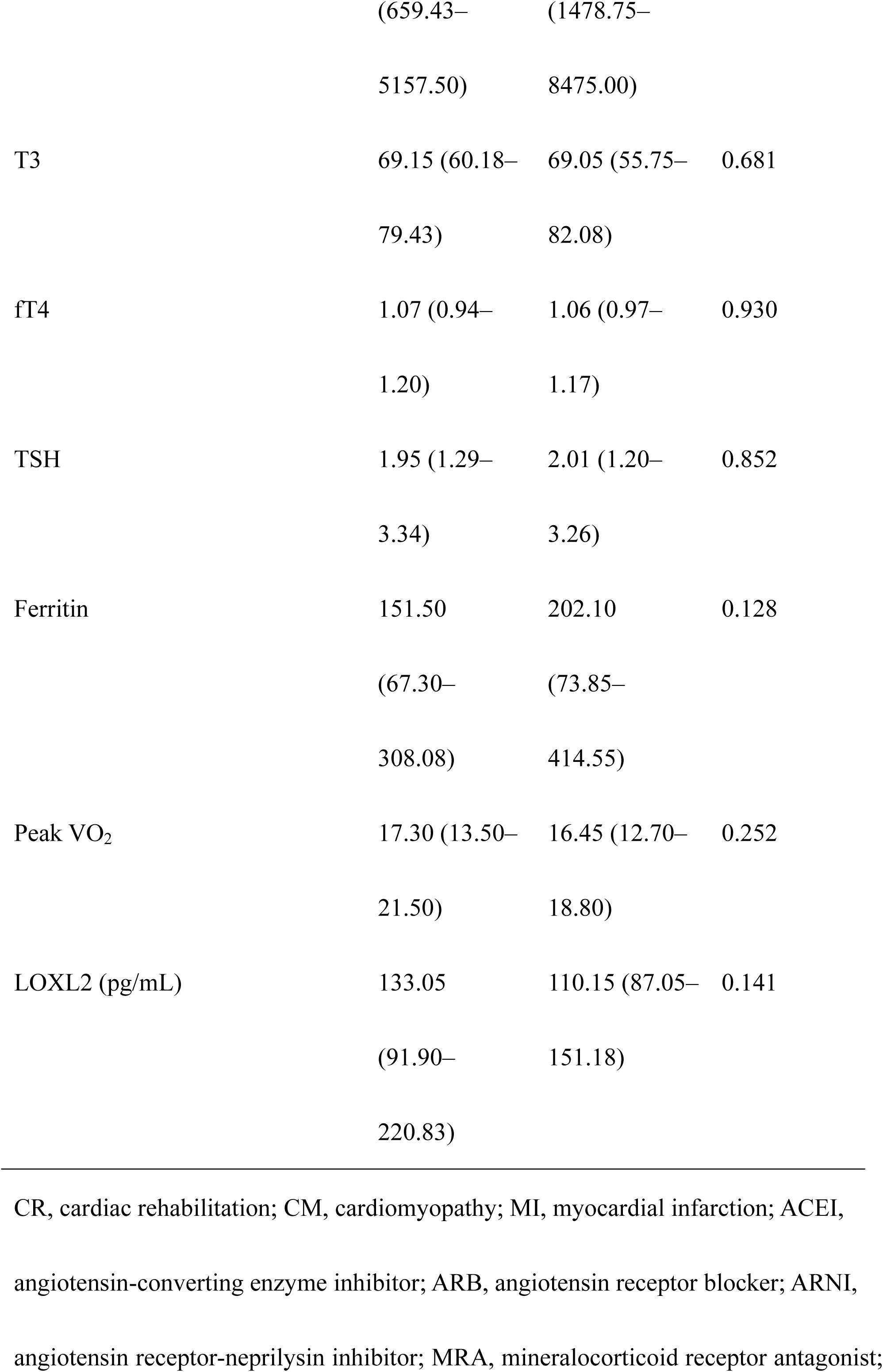
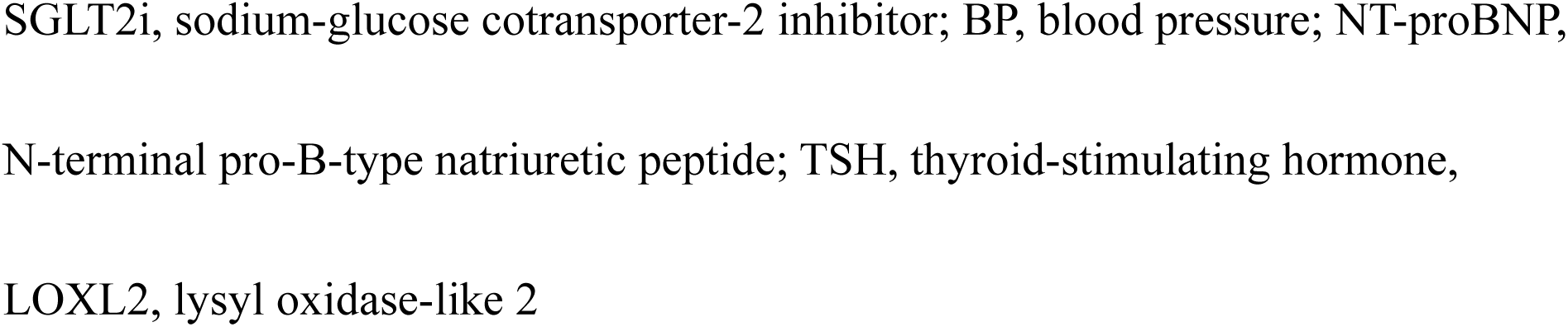
Demographic data of the study population.

**Table 2.** Demographic data for the study population after 1:1 propensity score matching.

| **Demographic characteristics** | **Early CR** | **Non-CR** | **Standardised** |
| --- | --- | --- | --- |
|  | **(*n*=33)** | **(*n*=33)** | **mean difference** |
|  | ***n* (% *N*) or** | ***n* (% *N*) or** |  |
|  | **mean (SD)** | **mean (SD)** |  |
| Age, median (range) | 52.67 (11.35) | 51.82<br>(13.93) | 0.072 |
| Sex, male | 26 (78.8%) | 27 (81.8%) | 0.076 |
| Ischaemic CM | 24.0 (72.7%) | 24.0 (72.7%) | <0.001 |
| Diabetes mellitus | 11.0 (33.3%) | 11.0 (33.3%) | <0.001 |
| Hypertension | 20.0 (60.6%) | 18.0 (54.5%) | 0.123 |
| Hyperlipidaemia | 8.0 (24.2%) | 7.0 (21.2%) | 0.072 |
| Previous MI | 2.0 (6.1%) | 1.0 (3.0%) | 0.146 |
| Atrial fibrillation | 2.0 (6.1%) | 2.0 (6.1%) | <0.001 |
| Body mass index (kg/m<sup>2</sup>), mean<br>(SD) | 26.50 (5.73) | 26.27 (6.29) | 0.039 |
| GDMT (≥3) | 21.0 (63.6%) | 19.0 (57.6%) | 0.124 |
| Estimated glomerular filtration | 72.87 | 75.22 | 0.091 |
| rate, mean (SD) | (25.21) | (26.17) |  |
| LOXL2 (pg/mL), mean (SD) | 152.09 | 137.12 | 0.202 |
|  | (77.64) | (70.07) |  |

CR, cardiac rehabilitation; CM, cardiomyopathy; MI, myocardial infarction; SD, standard deviation; LOXL2, lysyl oxidase-like 2

### Clinical outcomes

After PSM, the patients were evenly divided into the early CR and non-CR groups (*Table 2*). The all-cause mortality rate was significantly lower in the early CR group (0% vs. 21.2%, *p*=0.011) (*Table 3A*). No significant differences in the secondary endpoints, which included rehospitalisation for HF (*p*=0.269), improvement in KCCQ-12 scores at 6 months (*p*=0.387), and improvement in KCCQ-12 scores at 12 months (*p*=0.934), were identified between the groups (*Table 3A*).

**Table 3.**
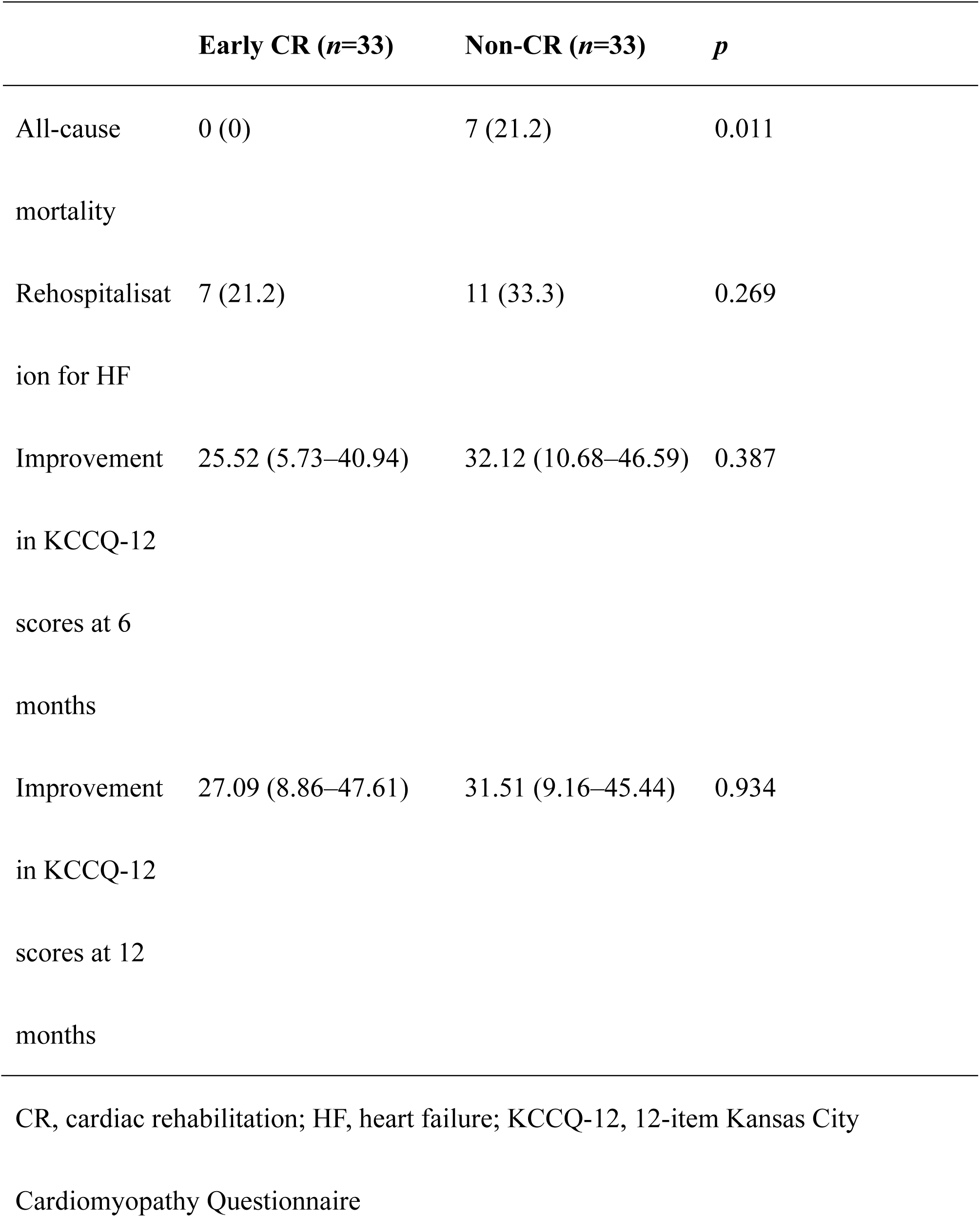
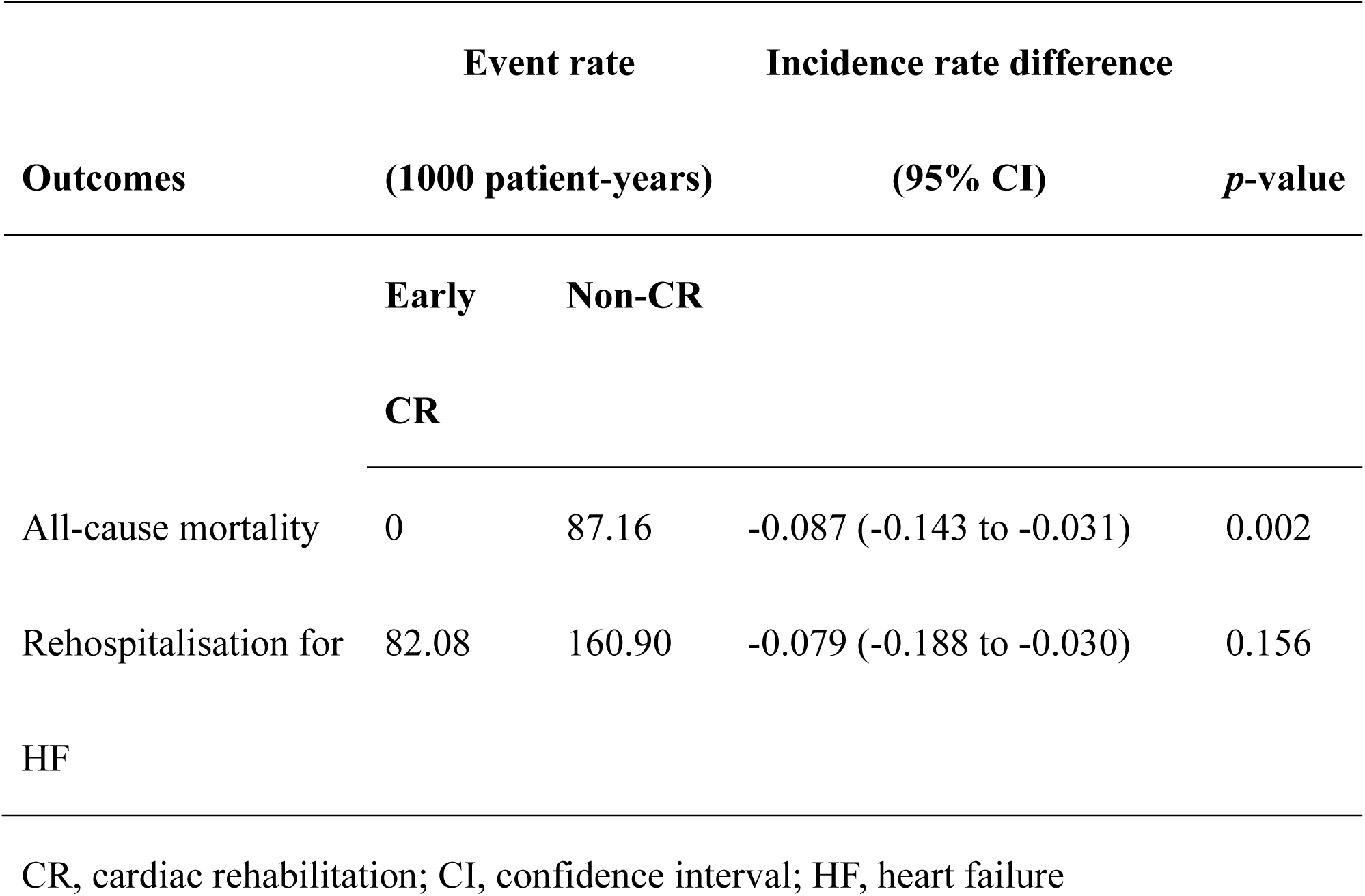
Clinical outcomes in patients with or without rehabilitation (propensity score-matched controls) Table 3A Primary and secondary endpoints, Table 3B Event rate and incidence rate difference

During a median follow-up period of 2.85 years (IQR: 2.33–3.83), patients who underwent CR showed a lower incidence rate of all-cause mortality (0 vs. 87.16 events per 1000 patient-years, rate difference: −0.087 [95% CI: −0.143 to −0.031], *p*=0.002) (*Table 3B*). However, the beneficial effect of CR on rehospitalisation for HF did not reach statistical significance (82.08 vs. 160.90 per 1000 patient-years, rate difference: −0.079 [95% CI: −0.188 to −0.030], *p*=0.156) (*Table 3B*). Patient survival rates according to participation in CR were examined using Kaplan–Meier curves, and *p*-values for differences between the early CR and non-CR groups were calculated using the log-rank test. The log-rank test indicated that CR was associated with a significantly reduced incidence of all-cause mortality (*p*=0.004) (*Figure 2A*); conversely, CR was not associated with a lower incidence of rehospitalisation for HF (*p*=0.264) (*Figure 2B*).

**Figure 2.**
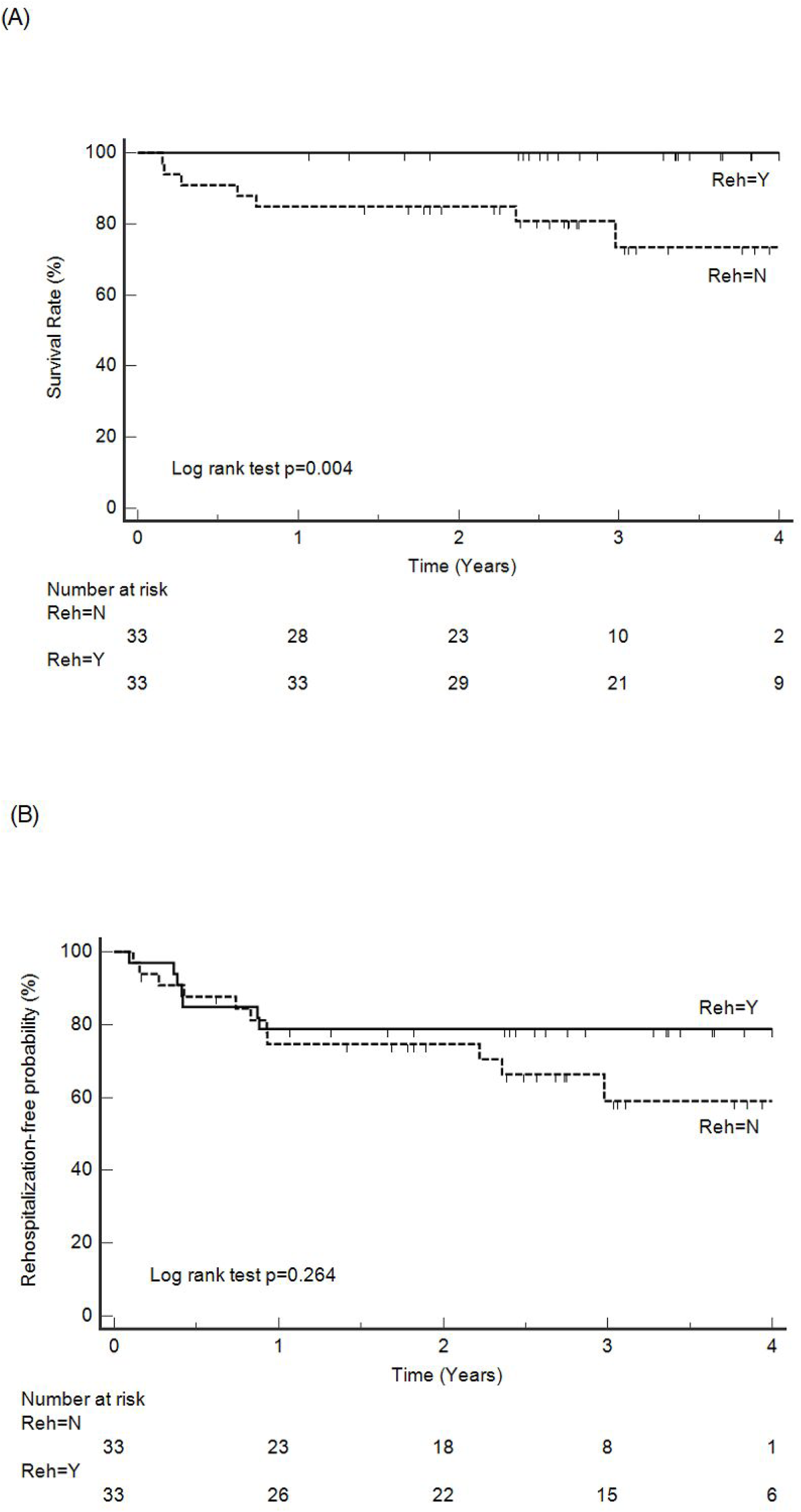
Cumulative incidence curves for (A) all-cause mortality and (B) rehospitalisation for HF in a propensity score-matched cohort. HF, heart failure Reh, Rehabilitation

Patients with higher LOXL2 levels (>200 pg/mL) at admission benefited from CR (0 vs. 172.3 events per 1000 patient-years, rate difference: −0.172 [95% CI: −0.299 to −0.046], *p*=0.008) (*Table 4*). However, the benefits of CR in patients with lower LOXL2 levels (0–100 pg/mL and >100 to ≤200 pg/mL) were not statistically significant (*p*=0.357 and *p*=0.068, respectively) (*Table 4*).

**Table 4.** Effects of CR on all-cause mortality according to LOXL2 level at admission.

|  | **Even rate** | **Incidence rate difference** |
| --- | --- | --- |

| Subgroups (*n*) | (1000 |  |  | *p*-value |
| --- | --- | --- | --- | --- |
|  | patient-years) |  |  |  |
|  | Early | Non-CR |  |  |
|  | CR |  |  |  |
| LOXL2 level: 0–100 pg/mL (*n*=61) | 0 | 31.29 | -0.031 (-0.098 to 0.035) | 0.357 |
| LOXL2 level: >100, £200 pg/mL (*n*=70) | 0 | 80.74 | -0.081 (-0.168 to 0.006) | 0.068 |
| LOXL2 level: >200 pg/mL (*n*=31) | 0 | 172.3 | -0.172 (-0.299 to -0.046) | 0.008 |

CR, cardiac rehabilitation; LOXL2, lysyl oxidase-like 2

During the CR period, no exercise-related deaths or major adverse events (e.g., arrhythmias, falls) occurred. Furthermore, no significant cardiovascular events associated with the home-based exercise program were reported. Additionally, exercise did not lead to the worsening of HF symptoms that would require hospitalisation.

## Discussion

This prospective cohort study revealed that the early exercise-based CR program initiated within 6 weeks of acute HF admission significantly reduced all-cause mortality over a nearly 3-year follow-up period, compared with that in the non-CR group. Notably, patients with more severe cardiac fibrosis derived significant benefits from the CR program. To our knowledge, this is the first prospective study to demonstrate that an early exercise-based CR program with a continued home-based CR program can reduce long-term all-cause mortality in patients discharged after an acute HF episode.

More than half of patients readmitted for HF within 30 days do not consult a physician or nursing specialist^12^, and those who do not receive outpatient follow-up within 7 days have the highest 30-day readmission rate^13^. The HFDMP, led by HF nursing specialists, has been shown to improve medication adherence and risk factor modification, effectively reducing the readmission rate^9,14^. In our study, both groups participated in an HFDMP led by an HF nursing specialist, which contributed to reduced readmissions in both groups. However, only the early CR group participated in the exercise-based and centre-based phase II CR program. This study demonstrated that integrating an early exercise program into the HFDMP, initiated within 6 weeks after discharge, could further significantly reduce all-cause mortality over a nearly 3-year follow-up period.

Early implementation of an exercise training program to prevent further physical deconditioning is considered beneficial for patients with HF. In the REHAB-HF trial, an exercise program was initiated early during hospitalisation; nevertheless, the REHAB-HF trial was insufficiently powered to evaluate clinical outcomes^5^. Moreover, the mortality rate appeared to increase in patients with HFrEF. A systematic review and meta-analysis of 13 RCTs investigating the effects of early exercise-based CR in patients with acute HF revealed that early exercise improved the exercise capacity (measured using the 6-minute walk distance and SPPB scores), quality of life (assessed using the Minnesota Living with Heart Failure Questionnaire and the KCCQ), and activities of daily living and reduced the all-cause readmission rate. However, no significant differences in LVEF, HF-related readmission rate, or all-cause mortality were observed^15^. Clinical guidelines recommend early ambulation and exercise training following acute cardiovascular events, such as acute myocardial infarction ^16^ and hospitalisation for acute HF^17^. A retrospective, multicentre, nationwide registry involving 10,473 patients from Japan demonstrated an association between early exercise intervention and a lower composite outcome (291 vs. 327 events per 1000 patient-years, rate ratio: 0.890 [95% CI: 0.830–0.954], *p*=0.001)^18^. Nonetheless, the incidence of cardiovascular mortality did not significantly differ between the CR and non-CR groups (rate ratio: 0.925 [95% CI: 0.824–1.038], *p*=0.18).

Additionally, most studies and meta-analyses have not found significant changes in all-cause mortality during a short follow-up period (<6 months)^19,20^. A recent large retrospective cohort study conducted within a global federated health research network in the United States indicated an association between exercise-based CR and a 42% reduction in the odds of all-cause mortality (odds ratio: 0.58, 95% CI: 0.54– 0.62) at a 2-year follow-up in 40,364 patients diagnosed with chronic HF^21^. Another multicentre, prospective cohort study with PSM involving 626 patients (313 pairs) reported that CR was associated with a reduced risk of all-cause mortality (HR: 0.53, 95% CI: 0.30–0.95, *p*=0.032) over a 2-year follow-up period. However, that multicentre study also revealed that regular CR did not confer survival benefits beyond 6 months after discharge^22^. Our prospective study, which incorporated an extended follow-up period and a home-based CR program, provides compelling evidence that supports the significant reduction in all-cause mortality achieved through early exercise-based CR initiation for acute HF following discharge.

Patients with HF accompanied by cardiac fibrosis experience considerably worse outcomes^6,23^, and few studies have demonstrated substantial benefits from either pharmacological treatments or non-pharmacological management in human participants^7,8,24^. There remains an unmet need to target maladaptive left ventricular tissue remodelling and fibrosis to improve the systolic and diastolic function of the myocardium^25^. Cardiac fibrosis not only disrupts the electromechanical coordination of cardiomyocytes, reducing systolic function^26^, but also increases ventricular stiffness, impairing diastolic function^27^. LOXL2 is upregulated in the interstitial tissue of failing hearts in both mice and humans. Elevated LOXL2 expression promotes transforming growth factor (TGF)-β2 production, inducing myofibroblast formation and migration, causing enhanced collagen deposition and crosslinking in hypertrophic regions of the stressed heart^28^. These pathological processes contribute to the development of cardiac fibrosis, ultimately resulting in myocardial dysfunction.

LOXL2 levels are strongly correlated with cardiac function and HF biomarkers^29^. Targeting LOXL2 in HF therapy is supported by findings from cell-based assays, pharmacological studies, mouse genetic models, and clinical evidence in patients with HF^30^. To our knowledge, this is the first study to demonstrate that exercise-based CR provides significant benefits to patients with HF accompanied by more severe cardiac fibrosis.

The beneficial effects of exercise-based CR may be attributed to two key mechanisms: pathophysiological mechanisms and psychosocial mechanisms^31^. With respect to pathophysiological mechanisms, exercise-based CR may contribute to the modification of cardiovascular risk factors, induction of ischaemic preconditioning, enhancement of cardiac electrical stability, improvement of myocardial oxygen supply, and better endothelial function. From a psychosocial perspective, exercise-based CR may enhance psychoneurological function, provide protection against depression and other psychological disorders, and facilitate behavioural changes beneficial for HF management^31^. Exercise enhances cardiopulmonary function, thereby improving patient survival. Potential mechanisms include an increased central cardiac output, enhanced peripheral oxygen consumption, better musculoskeletal function^32^, and improved vascular structure and function^33^. Exercise may also provide cardioprotective effects against cardiac fibrosis. The mechanism by which exercise mitigates cardiac fibrosis may involve promoting the secretion of cardioprotective exerkines, inhibiting systemic overactivation of the sympathetic nervous system and the renin–angiotensin system, attenuating oxidative stress and inflammatory responses, and modulating metabolism and noncoding RNA expression^34^. Aerobic exercise has been demonstrated by various animal studies to provide potential benefits in mitigating cardiac fibrosis. In mice with vitamin D deficiency, exercise was shown to alleviate cardiac fibrosis, an effect associated with increased expression of the vitamin D receptor, inhibition of the TGF-β1/mothers against decapentaplegic homolog 2/3 (Smad2/3) signalling pathway, and reduced expression of profibrotic and inflammatory factors^35^. An 8-week aerobic exercise program has been shown to reduce cardiac fibrosis by downregulating angiotensin type-1 receptor expression in hypertensive ovariectomised rats^36^. Experimental studies have shown that exercise stimulates the release of plasma exosomes from endothelial progenitor cells, which are enriched with microRNAs, such as miR-126^37^. These exosomes exhibit cardioprotective properties by activating cardiac fibroblasts, enhancing the expression of endothelial cell-specific markers, and suppressing proteins associated with fibrosis^38^. Specifically, miR-126 reduces cardiac fibrosis by inhibiting the expression of TGF-β, Smad2, and Smad3^39^. The exercise-induced release of exosomes, which mitigates cardiac fibrosis, provides a novel mechanism underlying the therapeutic benefits of CR.

The low participation rate in CR, particularly after acute HF, is a global issue. In this study, although we enrolled 162 patients, only 21% (34 patients) participated in outpatient CR. Similarly, data from the Global Federated Health Research Network in the United States showed that only 1.6% patients with HF engaged in CR^21^. A multicentre, retrospective cohort study in Japan reported a participation rate of 26% (862/3277) for patients hospitalised with acute HF^40^. On the basis of the findings of this study, particularly the effectiveness of home-based CR, clinicians should make every effort to encourage their patients to engage in early post-discharge CR and exercise therapy to improve their outcomes.

This study has some limitations. First, while our study provides real-world data, it is a small cohort study conducted at a single medical centre with an open-label design. Second, the decision to participate in CR was based on patients’ personal choice, which might have led to a greater likelihood for patients with better overall conditions to opt for CR. Despite the use of PSM analysis to make the evaluation more objective, this selection bias might still have influenced the outcomes. Third, the outbreak of the COVID-19 pandemic occurred during our enrolment period, and home-based CR for the CR group was implemented. However, adherence to home-based CR could not be fully monitored. Furthermore, although we expended efforts to track the patients using heart rate variability and video consultations, this was not as precise as centre-based CR. Nevertheless, this experience highlights that home-based CR could be a potential area for future development. Fourth, the effectiveness of CR may follow a dose–response relationship, which was not evaluated in this study. Fifth, comprehensive follow-up assessments of patients’ cardiopulmonary exercise tests, which could serve as an important prognostic indicator, were not performed. Lastly, our study did not analyse psychological factors, including depression and cognitive function, which are critical components of a multidisciplinary CR program^10^. Future large-scale studies incorporating these factors should be conducted for a more comprehensive evaluation.

In conclusion, the current study revealed that the combination of early post-discharge exercise therapy and home-based CR was associated with reduced all-cause mortality in patients with acute HF who participated in the HFDMP. Individuals with more advanced cardiac fibrosis seemed to derive greater benefits from exercise-based CR. Our findings suggest that exercise therapy could be a potential treatment strategy for targeting elevated LOXL2 levels in patients with HF. This study underscores the importance of early CR and the need for large-scale studies to validate these findings.

## Data Availability

The data underlying this article will be shared on reasonable request to the corresponding author.

## Acknowledgements

We would like to thank Hsin-Yi Chien, Chih-Yun Lin, and the Biostatistics Center at Kaohsiung Chang Gung Memorial Hospital for their guidance with the statistical analyses.

## Authors’ contributions

SM Chen: Conceptualization, Writing-original draft, review and editing. LY Wang: Design exercise program. HY Hsiao, YC Zheng, PJ Wu: Investigation and Data curation. CL Wei: Coordinator of disease management program. CJ Chen: Supervision, Validation. CL Hang: Supervision, Validation. S Leu: LOXL2 analysis. YL Chen: Supervision.

## References

1. Hung CL, Chao TF, Tsai CT, Liao JN, Lim SS, Tuan TC, Chen TJ, Chan YH, Chen SA, Chiang CE. Prevalence, Incidence, Lifetime Risks, and Outcomes of Heart Failure in Asia: A Nationwide Report. JACC Heart Fail. 2023;11:1454–1456. doi: 10.1016/j.jchf.2022.07.012

2. Bjarnason-Wehrens B, Nebel R, Jensen K, Hackbusch M, Grilli M, Gielen S, Schwaab B, Rauch B, German Society of Cardiovascular P, Rehabilitation. Exercise-based cardiac rehabilitation in patients with reduced left ventricular ejection fraction: The Cardiac Rehabilitation Outcome Study in Heart Failure (CROS-HF): A systematic review and meta-analysis. Eur J Prev Cardiol. 2020;27:929–952. doi: 10.1177/2047487319854140

3. O’Connor CM, Whellan DJ, Lee KL, Keteyian SJ, Cooper LS, Ellis SJ, Leifer ES, Kraus WE, Kitzman DW, Blumenthal JA, et al. Efficacy and safety of exercise training in patients with chronic heart failure: HF-ACTION randomized controlled trial. JAMA. 2009;301:1439–1450. doi: 10.1001/jama.2009.454

4. Chen SM, Wang LY, Liaw MY, Wu MK, Wu PJ, Wei CL, Chen AN, Su TL, Chang JK, Yang TH, et al. Outcomes With Multidisciplinary Cardiac Rehabilitation in Post-acute Systolic Heart Failure Patients-A Retrospective Propensity Score-Matched Study. Front Cardiovasc Med. 2022;9:763217. doi: 10.3389/fcvm.2022.763217

5. Kitzman DW, Whellan DJ, Duncan P, Pastva AM, Mentz RJ, Reeves GR, Nelson MB, Chen H, Upadhya B, Reed SD, et al. Physical Rehabilitation for Older Patients Hospitalized for Heart Failure. N Engl J Med. 2021;385:203–216. doi: 10.1056/NEJMoa2026141

6. Heymans S, Gonzalez A, Pizard A, Papageorgiou AP, Lopez-Andres N, Jaisser F, Thum T, Zannad F, Diez J. Searching for new mechanisms of myocardial fibrosis with diagnostic and/or therapeutic potential. Eur J Heart Fail. 2015;17:764–771. doi: 10.1002/ejhf.312

7. Gyongyosi M, Winkler J, Ramos I, Do QT, Firat H, McDonald K, Gonzalez A, Thum T, Diez J, Jaisser F, et al. Myocardial fibrosis: biomedical research from bench to bedside. Eur J Heart Fail. 2017;19:177–191. doi: 10.1002/ejhf.696

8. de Boer RA, De Keulenaer G, Bauersachs J, Brutsaert D, Cleland JG, Diez J, Du XJ, Ford P, Heinzel FR, Lipson KE, et al. Towards better definition, quantification and treatment of fibrosis in heart failure. A scientific roadmap by the Committee of Translational Research of the Heart Failure Association (HFA) of the European Society of Cardiology. Eur J Heart Fail. 2019;21:272–285. doi: 10.1002/ejhf.1406

9. Chen SM, Fang YN, Wang LY, Wu MK, Wu PJ, Yang TH, Chen YL, Hang CL. Impact of multi-disciplinary treatment strategy on systolic heart failure outcome. BMC Cardiovasc Disord. 2019;19:220. doi: 10.1186/s12872-019-1214-0

10. Chen SM, Wu MK, Chen C, Wang LY, Guo NW, Wei CL, Zheng YC, Hsiao HY, Wu PJ, Chen YL, et al. Benefit of cardiac rehabilitation in acute heart failure patients with cognitive impairment. Heliyon. 2024;10:e30493. doi: 10.1016/j.heliyon.2024.e30493

11. Chen SM, Wang LY, Wu PJ, Liaw MY, Chen YL, Chen AN, Tsai TH, Hang CL, Lin MC. The Interrelationship between Ventilatory Inefficiency and Left Ventricular Ejection Fraction in Terms of Cardiovascular Outcomes in Heart Failure Outpatients. Diagnostics (Basel). 2020;10. doi: 10.3390/diagnostics10070469

12. Soundarraj D, Singh V, Satija V, Thakur RK. Containing the Cost of Heart Failure Management: A Focus on Reducing Readmissions. Heart Fail Clin. 2017;13:21–28. doi: 10.1016/j.hfc.2016.07.002

13. Hernandez AF, Greiner MA, Fonarow GC, Hammill BG, Heidenreich PA, Yancy CW, Peterson ED, Curtis LH. Relationship Between Early Physician Follow-up and 30-Day Readmission Among Medicare Beneficiaries Hospitalized for Heart Failure. JAMA. 2010;303:1716–1722. doi: 10.1001/jama.2010.533

14. Ruppert L, Williams S, Bhatia-Patel S, Fabrizio C, Rakita V, Hagy A, Hamilton J, Fontana S, Hamad E. Heart Failure Disease Management Program Decreases 30-day Re-admissions & Improves Heart Failure Quality Metrics. Journal of Cardiac Failure. 2024;30:264. doi: 10.1016/j.cardfail.2023.10.349

15. Meng Y, Zhuge W, Huang H, Zhang T, Ge X. The effects of early exercise on cardiac rehabilitation-related outcome in acute heart failure patients: A systematic review and meta-analysis. Int J Nurs Stud. 2022;130:104237. doi: 10.1016/j.ijnurstu.2022.104237

16. Ibanez B, James S, Agewall S, Antunes MJ, Bucciarelli-Ducci C, Bueno H, Caforio ALP, Crea F, Goudevenos JA, Halvorsen S, et al. 2017 ESC Guidelines for the management of acute myocardial infarction in patients presenting with ST-segment elevation: The Task Force for the management of acute myocardial infarction in patients presenting with ST-segment elevation of the European Society of Cardiology (ESC). Eur Heart J. 2018;39:119–177. doi: 10.1093/eurheartj/ehx393

17. Heidenreich PA, Bozkurt B, Aguilar D, Allen LA, Byun JJ, Colvin MM, Deswal A, Drazner MH, Dunlay SM, Evers LR, et al. 2022 AHA/ACC/HFSA Guideline for the Management of Heart Failure: Executive Summary: A Report of the American College of Cardiology/American Heart Association Joint Committee on Clinical Practice Guidelines. Circulation. 2022;145:e876–e894. doi: 10.1161/CIR.0000000000001062

18. Enzan N, Matsushima S, Kaku H, Tohyama T, Nezu T, Higuchi T, Nagatomi Y, Fujino T, Hashimoto T, Ide T, et al. Propensity-Matched Study of Early Cardiac Rehabilitation in Patients With Acute Decompensated Heart Failure. Circ Heart Fail. 2023; 16:e010320. doi: 10.1161/CIRCHEARTFAILURE.122.010320

19. McKelvie RS. Exercise training in patients with heart failure: clinical outcomes, safety, and indications. Heart Fail Rev. 2008;13:3–11. doi: 10.1007/s10741-007-9052-z

20. Molloy C, Long L, Mordi IR, Bridges C, Sagar VA, Davies EJ, Coats AJ, Dalal H, Rees K, Singh SJ, et al. Exercise-based cardiac rehabilitation for adults with heart failure. Cochrane Database Syst Rev. 2024;3:CD003331. doi: 10.1002/14651858.CD003331.pub6

21. Buckley BJR, Harrison SL, Fazio-Eynullayeva E, Underhill P, Sankaranarayanan R, Wright DJ, Thijssen DHJ, Lip GYH. Cardiac rehabilitation and all-cause mortality in patients with heart failure: a retrospective cohort study. Eur J Prev Cardiol. 2021;28:1704–1710. doi: 10.1093/eurjpc/zwab035

22. Adachi T, Iritani N, Kamiya K, Iwatsu K, Kamisaka K, Iida Y, Yamada S, collaborators F. Prognostic Effects of Cardiac Rehabilitation in Patients With Heart Failure (from a Multicenter Prospective Cohort Study). Am J Cardiol. 2022;164:79–85. doi: 10.1016/j.amjcard.2021.10.038

23. Schelbert EB, Piehler KM, Zareba KM, Moon JC, Ugander M, Messroghli DR, Valeti US, Chang CC, Shroff SG, Diez J, et al. Myocardial Fibrosis Quantified by Extracellular Volume Is Associated With Subsequent Hospitalization for Heart Failure, Death, or Both Across the Spectrum of Ejection Fraction and Heart Failure Stage. J Am Heart Assoc. 2015;4. doi: 10.1161/JAHA.115.002613

24. Clarke SA, Richardson WJ, Holmes JW. Modifying the mechanics of healing infarcts: Is better the enemy of good? J Mol Cell Cardiol. 2016;93:115–124. doi: 10.1016/j.yjmcc.2015.11.028

25. Bonanni A, Vinci R, d’Aiello A, Grimaldi MC, Di Sario M, Tarquini D, Proto L, Severino A, Pedicino D, Liuzzo G. Targeting Collagen Pathways as an HFpEF Therapeutic Strategy. J Clin Med. 2023;12. doi: 10.3390/jcm12185862

26. Humeres C, Frangogiannis NG. Fibroblasts in the Infarcted, Remodeling, and Failing Heart. JACC Basic Transl Sci. 2019;4:449–467. doi: 10.1016/j.jacbts.2019.02.006

27. Homsi R, Luetkens JA, Skowasch D, Pizarro C, Sprinkart AM, Gieseke J, Meyer Zur Heide Gen Meyer-Arend J, Schild HH, Naehle CP. Left Ventricular Myocardial Fibrosis, Atrophy, and Impaired Contractility in Patients With Pulmonary Arterial Hypertension and a Preserved Left Ventricular Function: A Cardiac Magnetic Resonance Study. J Thorac Imaging. 2017;32:36–42. doi: 10.1097/RTI.0000000000000248

28. Yang J, Savvatis K, Kang JS, Fan P, Zhong H, Schwartz K, Barry V, Mikels-Vigdal A, Karpinski S, Kornyeyev D, et al. Targeting LOXL2 for cardiac interstitial fibrosis and heart failure treatment. Nat Commun. 2016;7:13710. doi: 10.1038/ncomms13710

29. Rodriguez C, Martinez-Gonzalez J. The Role of Lysyl Oxidase Enzymes in Cardiac Function and Remodeling. Cells. 2019;8. doi: 10.3390/cells8121483

30. Poe A, Martinez Yus M, Wang H, Santhanam L. Lysyl oxidase like-2 in fibrosis and cardiovascular disease. Am J Physiol Cell Physiol. 2023;325:C694–C707. doi: 10.1152/ajpcell.00176.2023

31. Chen KC, Hsu CN, Wu CH, Lin KL, Chen SM, Lee Y, Hsu CY, Hsu CW, Huang CY, Huang SH, et al. 2023 TAMIS/TSOC/TACVPR Consensus Statement for Patients with Acute Myocardial Infarction Rehabilitation. Acta Cardiol Sin. 2023;39:783–806. doi: 10.6515/ACS.202311_39(6).20230921A

32. van Tol BA, Huijsmans RJ, Kroon DW, Schothorst M, Kwakkel G. Effects of exercise training on cardiac performance, exercise capacity and quality of life in patients with heart failure: a meta-analysis. Eur J Heart Fail. 2006;8:841–850. doi: 10.1016/j.ejheart.2006.02.013

33. Green DJ, Hopman MT, Padilla J, Laughlin MH, Thijssen DH. Vascular Adaptation to Exercise in Humans: Role of Hemodynamic Stimuli. Physiol Rev. 2017;97:495–528. doi: 10.1152/physrev.00014.2016

34. Wang K, Deng Y, Xiao H. Exercise and cardiac fibrosis. Current Opinion in Physiology. 2023;31:100630. doi: 10.1016/j.cophys.2022.100630

35. Cui X, Wang K, Zhang J, Cao ZB. Aerobic Exercise Ameliorates Myocardial Fibrosis via Affecting Vitamin D Receptor and Transforming Growth Factor-beta1 Signaling in Vitamin D-Deficient Mice. Nutrients. 2023;15. doi: 10.3390/nu15030741

36. Lin Y-Y, Hong Y, Zhou M-C, Huang H-L, Shyu W-C, Chen J-S, Ting H, Cheng Y-J, Yang A-L, Lee S-D. Exercise training attenuates cardiac inflammation and fibrosis in hypertensive ovariectomized rats. Journal of Applied Physiology. 2020;128:1033–1043. doi: 10.1152/japplphysiol.00844.2019

37. Ma C, Wang J, Liu H, Chen Y, Ma X, Chen S, Chen Y, Bihl JI, Yang YI. Moderate Exercise Enhances Endothelial Progenitor Cell Exosomes Release and Function. Med Sci Sports Exerc. 2018;50:2024–2032. doi: 10.1249/MSS.0000000000001672

38. Ke X, Yang D, Liang J, Wang X, Wu S, Wang X, Hu C. Human Endothelial Progenitor Cell-Derived Exosomes Increase Proliferation and Angiogenesis in Cardiac Fibroblasts by Promoting the Mesenchymal-Endothelial Transition and Reducing High Mobility Group Box 1 Protein B1 Expression. DNA Cell Biol. 2017;36:1018–1028. doi: 10.1089/dna.2017.3836

39. Fu G, Wang Z, Hu S. Exercise improves cardiac fibrosis by stimulating the release of endothelial progenitor cell-derived exosomes and upregulating miR-126 expression. Front Cardiovasc Med. 2024;11:1323329. doi: 10.3389/fcvm.2024.1323329

40. Kamiya K, Sato Y, Takahashi T, Tsuchihashi-Makaya M, Kotooka N, Ikegame T, Takura T, Yamamoto T, Nagayama M, Goto Y, et al. Multidisciplinary Cardiac Rehabilitation and Long-Term Prognosis in Patients With Heart Failure. Circ Heart Fail. 2020;13:e006798. doi: 10.1161/CIRCHEARTFAILURE.119.006798

